# Healthcare associated infection risk analysis at the obstetrics and gynecology department of a referral hospital in Cameroon

**DOI:** 10.1101/2025.05.03.25326934

**Authors:** Fabrice Zobel Lekeumo Cheuyem, Emilia Enjema Lyonga, Innocent Takougang

**Author notes:** Corresponding author’s address: Fabrice Zobel Lekeumo Cheuyem, Department of Public Health, Faculty of Medicine & Biomedical Sciences, The University of Yaounde I, Yaounde, Cameroon.

## Abstract

**Background:** Healthcare-associated infections (HAIs) are a major problem in healthcare facilities. In Cameroon, maternal and neonatal mortality remain a concern. The underlying determinants include shortcomings in the quality of care, inadequate infrastructure and inconsistent application of infection control and prevention (ICP) measures. The objective of this study was to identify risks that increase the likelihood of HAIs in the obstetrics-gynecology unit of a referral hospital of Yaoundé.

**Methods:** A descriptive cross-sectional study was conducted in the obstetrics-gynecology department of referral hospital of Yaoundé from April to July 2024. Exposure data were collected using a self-administered questionnaire with a scoring grid to assess the level of implementation of the ICP framework. Samples were collected from the ward environment for identification of the bacterial flora of the care environment and antibiotic susceptibility testing.

**Results:** A total of 41 healthcare workers were enrolled in the study. Participants were predominantly female (78%) and aged 20 to 57 (median of 30) years. Hand hygiene knowledge of was average, with a median score of 60%. More than two-thirds of respondents (n=30; 73%) reported that they did not systematically practice hand hygiene before and after patient care. The most common reason for not practicing hand hygiene was the absence of a hand-washing site nearby healthcare post (57%). Face shields were the most reported unavailable equipment (81%). The overall score indicated that the obstetrics-gynecology department had a basic level of implementation of ICP interventions. Microbial analysis revealed that taps and trolleys were the most commonly soiled equipment, harboring bacterial flora of Staphylococcus aureus (36%) and other Staphylococcus spp. (24%). Proteus mirabilis (13%) and Klebsiella spp. (7%). The isolated bacterial strains showed varying degrees of resistance to antibiotics, including cephalosporin, methicillin and penicillin.

**Conclusions:** Suboptimal IPC implementation was observed in this obstetrics-gynecology setting. Comprehensive interventions are needed, including: strengthened IPC adherence, national antibiotic regulation, healthcare worker education, and establishment of antimicrobial resistance surveillance programs.

## Background

Healthcare-associated infections (HAIs) are common adverse events in healthcare settings [1]. HAIs include infections acquired during care in health facilities, either during admission or after discharge [1,2]. Among the 190 million annual hospital admissions, 9 million HAIs occur, resulting in 1 million fatalities [3]. HAIs contribute to the emergence of antimicrobial-resistant pathogens that complicate infection management and threaten patient safety [4]. Disparities exist in the burden of HAIs, with low- and middle-income countries disproportionately affected, where 63.5% of infections are caused by antimicrobial-resistant bacteria [5,6].

Cameroon has witnessed a significant increase in the number of women seeking antenatal care at health facilities in recent years [7]. Maternal (596 per 100,000 live births) and neonatal (25.6 per 1,000 live births) mortality remain high in the country [8,9]. Maternal mortality has been associated with sepsis, abortion, place of delivery, and quality of care during childbirth [10]. Deficiencies in the quality of delivery service contribute to puerperal infections (9-17%) and post-caesarean wound infections (2-32%) across primary, secondary, and tertiary health facilities [11–13].

Maternal and neonatal sepsis-related mortality highlights wider healthcare determinants and underlying quality of care issues, infrastructure constraints, inconsistent compliance with infection control measures, and diagnostic delays, all of which have been identified as major determinants of HAIs and their complications. Healthcare workers’ (HCWs) adherence to standard precautions and the scarcity of infection prevention specialists to support the implementation of infection control and prevention (ICP) measures in low-income countries are significant constraints to reducing infection risk and contribute to poor adherence to ICP measures during labor and delivery [14].

In a context of poor regulation of antibiotic use, an increase in the burden of multi-resistant bacteria has been observed, particularly in healthcare settings [15,16]. Literature on adherence to standard precautions, implementation of ICP, and bacterial carriage in Cameroonian health facilities is limited [16–18]. To support evidence-based decision-making and policy development, an analysis of the situation in health facilities providing maternal and newborn care was needed. The present study was designed to assess the risk of HAIs in the obstetrics and gynecology ward of a referral hospital in Yaoundé.

## Methods

The present investigation focused on HCWs and the care environment to determine the microbial flora and assess the risk of exposure. It was a descriptive cross-sectional study carried out from May to July 2024.

The study was conducted in Yaoundé, the capital city of Cameroon, a city of approximately 3.2 million inhabitants served by first, second, third, and fourth-level health facilities. The healthcare setting studied is ranked as a fourth-level referral center in the healthcare pyramid. It provides both short- and long-term specialized care to the population and plays a key role in the training of satellite HCWs [19,20].

It has a capacity of 138 beds and receives between 500 and 1,000 patients per week. In 2023, it recorded 35,945 outpatient consultations and 4,329 admissions, totaling 21,518 hospital days with an average of 5 hospital days per inpatient [19,21]. For the year 2023, the obstetrics-gynecology department reported approximately 300 deliveries, 8 deaths, 625 prenatal consultations, 77 cesarean sections, and managed 101 obstetric complications and 33 abortion complications [21].

### Assessment of exposure

All HCWs operating in the obstetrics-gynecology ward were invited to participate. This included residents, nurses, midwives/obstetric nurses, assistant nurses, and students. All consenting HCWs were included in the study.

A pre-tested questionnaire, adapted from the knowledge assessment tool developed by the WHO, was administered to HCWs [22,23]. The questionnaire included items related to demographic characteristics, hand hygiene, availability of personal protective equipment (PPE), waste management, and sterilization. The questionnaire was anonymous and self-administered. A standard form developed by the WHO was adapted to serve as a tool for assessing the ICP implementation framework in the care unit [24,25]. The ICP implementation framework assessment form was administered during interviews with the department’s key informants (sub-unit head nurses).

### Assessment of microbial flora of the care environment

A total of 30 samples were planned for collection during this study. These samples included medical equipment such as the delivery table, trolley, drip stand, nursing table, and surfaces (tap, door handle). A purposive sampling design was used. Ten samples were collected weekly for three weeks, divided into two phases of five samples, one day apart.

Surface and medical device samples were collected in the early morning hours using sterile swabs premoistened with sterile saline solution. Swabs were moistened with 0.9% sodium chloride solution and passed across the surface in parallel, tight stripes, then repeated perpendicularly. Swabs were placed in labeled cryotubes containing brain heart infusion broth and were immediately sent to the laboratory for bacteriological examination [16,26].

Upon arrival at the bacteriology laboratory, each cotton swab was aseptically transferred to a tube containing 2 mL of sterile physiological solution, from which 0.1-0.4 mL aliquots were pipetted into selective culture media (eosin-methylene blue agar for Gram-negative bacteria and mannitol salt agar for Staphylococcus spp.). The inoculated media were incubated at 37°C for 24-48 hours for specific pathogens. After incubation, bacterial isolates were identified using standard biochemical methods (Gram-negative bacilli were identified using reaction to glucose, lactose, citrate, H₂S, lactose, urea, indoles, and gas production).

Antibiotic susceptibility testing was performed using the Müller-Hinton agar disk diffusion technique [16]. Bacterial suspensions were prepared according to the 0.5 McFarland standard. Discs were placed on inoculated agar and incubated at 37°C ± 2°C for 18-24 hours. Selection and susceptibility of the antibiotic discs were performed according to the guidelines of the European Committee on Antimicrobial Susceptibility Testing (EUCAST) 2023 [27]. Detected Staphylococcus aureus strains were tested for methicillin resistance using a cefoxitin disc. Strains with an inhibition diameter <27 mm were considered methicillin-resistant S. aureus (MRSA) [26,28]. To minimize potential bias, we performed rigorous quality control on culture conditions, media preparation, antibiotic authenticity, equipment calibration, and reagent validation to ensure the reliability and accuracy of our findings.

A score of 1 was assigned for each correct response to the hand hygiene knowledge assessment. At the end of this assessment, the results were classified into three categories: good (≥75%), average (50-74%), and poor (<50%) [29]. The professional group variable included medical staff (residents), paramedical professionals (nurses, midwives/obstetric nurses, assistant nurses), and students (medical undergraduates). A score was assigned for each question on the ICP implementation framework assessment grid. Each of the eight components was scored out of 100, resulting in a total score of 800. Data were checked, coded, and analyzed using R Statistics version 4.3.3. Fisher’s exact probability test was used to compare proportions. Confidence intervals (CI) were estimated at 95%. A p-value of less than 0.05 was considered statistically significant.

The protocol was approved by the Institutional Research Ethics Committee (CIER) of the Faculty of Medicine and Biomedical Sciences of the University of Yaoundé I, and ethical approval was issued under number 1017/UYI/FMSB/VDRC/DAASR/CSD. Research approval was also obtained from the general management of the healthcare setting. Informed consent was obtained from the participants prior to their inclusion in the study. All procedures were carried out in accordance with the Declaration of Helsinki.

## Results

Of the 47 HCWs contacted, 46 (98%) agreed to take part in the study. A total of 41 HCWs returned fully completed questionnaires, representing a response rate of (89%).

### Socio-professional characteristics of study participants

Participants aged 20 to 57 years, with a median age of 30 [22; 47] years. They were predominantly female (78%) and had higher education (73.2%). Their median work experience was 3 [3; 19] years.

### Hand hygiene knowledge

Knowledge of hand hygiene was average, with a median score of 60% [53; 67]. Residents showed the best knowledge of hand hygiene (100% having average or good knowledge; p=0.296). The lowest knowledge levels were observed among midwives (29%) and younger participants, notably medical students (30%). In addition, those who reported having received specific training on hand hygiene were significantly (p=0.019) less knowledgeable on the subject (Table 1).

**Table 1.** Relationship between socioprofessional characteristics and hand hygiene knowledge among healthcare workers.

| Variable | Knowledge Assessment |  |  | Total | *p*-value<sup>1</sup> |
| --- | --- | --- | --- | --- | --- |
|  | *n* (%) |  |  |  |  |
|  | Poor | Average | Good |  |  |
|  | 9 (22) | 29 (71) | 3 (7) | 41 (100) |  |
| Age (in years) |  |  |  |  | 0.516 |
| □25 | 11 (73) | 4 (27) | 0 (0.0) | 15 |  |
| 25 - 44 | 11 (78) | 2 (14) | 1 (7) | 14 |  |
| 45 + | 7 (58) | 3 (25) | 2 (17) | 12 |  |
| Educational level |  |  |  |  | 0.147 |
| Secondary | 1 (9) | 8 (73) | 2 (18) | 11 |  |
| Higher | 8 (27) | 21 (70) | 1 (3) | 30 |  |
| Professional status |  |  |  |  | 0.296 |
| Assistant-nurse | 1 (33) | 1 (33) | 1 (34) | 3 |  |
| Medical student | 3 (30) | 7 (70) | 0 (0) | 10 |  |
| Nurse | 3 (23) | 9 (69) | 1 (8) | 13 |  |
| Resident | 0 (0) | 9 (75) | 1 (25) | 10 |  |
| Midwife/obstetric nurse | 2 (29) | 5 (71) | 0 (0) | 7 |  |
| Professional group |  |  |  |  | 0.430 |
| Student | 3 (30) | 7 (70) | 0 (0) | 10 |  |
| Medical | 0 (0) | 7 (88) | 1 (12) | 8 |  |
| Paramedical | 6 (26) | 15 (65) | 2 (9) | 23 |  |
| Professional experience (in years) |  |  |  |  | 0.170 |
| □10 | 17 (71) | 6 (25) | 1 (4) | 24 |  |
| 10 + | 12 (71) | 3 (18) | 2 (11) | 17 |  |
| Refresher course on ICP measures |  |  |  |  | >0.999 |
| No | 6 (22) | 19 (71) | 2 (7) | 27 |  |
| Yes | 3 (21) | 10 (72) | 1 (7) | 14 |  |
| Training received on hand hygiene |  |  |  |  | **0.019** |
| No | 1 (6) | 13 (76) | 3 (18) | 17 |  |
| Yes | 8 (33) | 16 (67) | 0 (0) | 24 |  |

<sup>1</sup>Fisher exact probability test; ICP: Infection Control and Prevention

Over two-thirds of respondents reported not systematically practicing hand hygiene before and after care (n=30; 73%). The main reasons mentioned included the absence of a hand-washing site nearby (57%), the perception that patient care was safe (30%) and high workload (20%) (Figure 1).

**Fig. 1.**
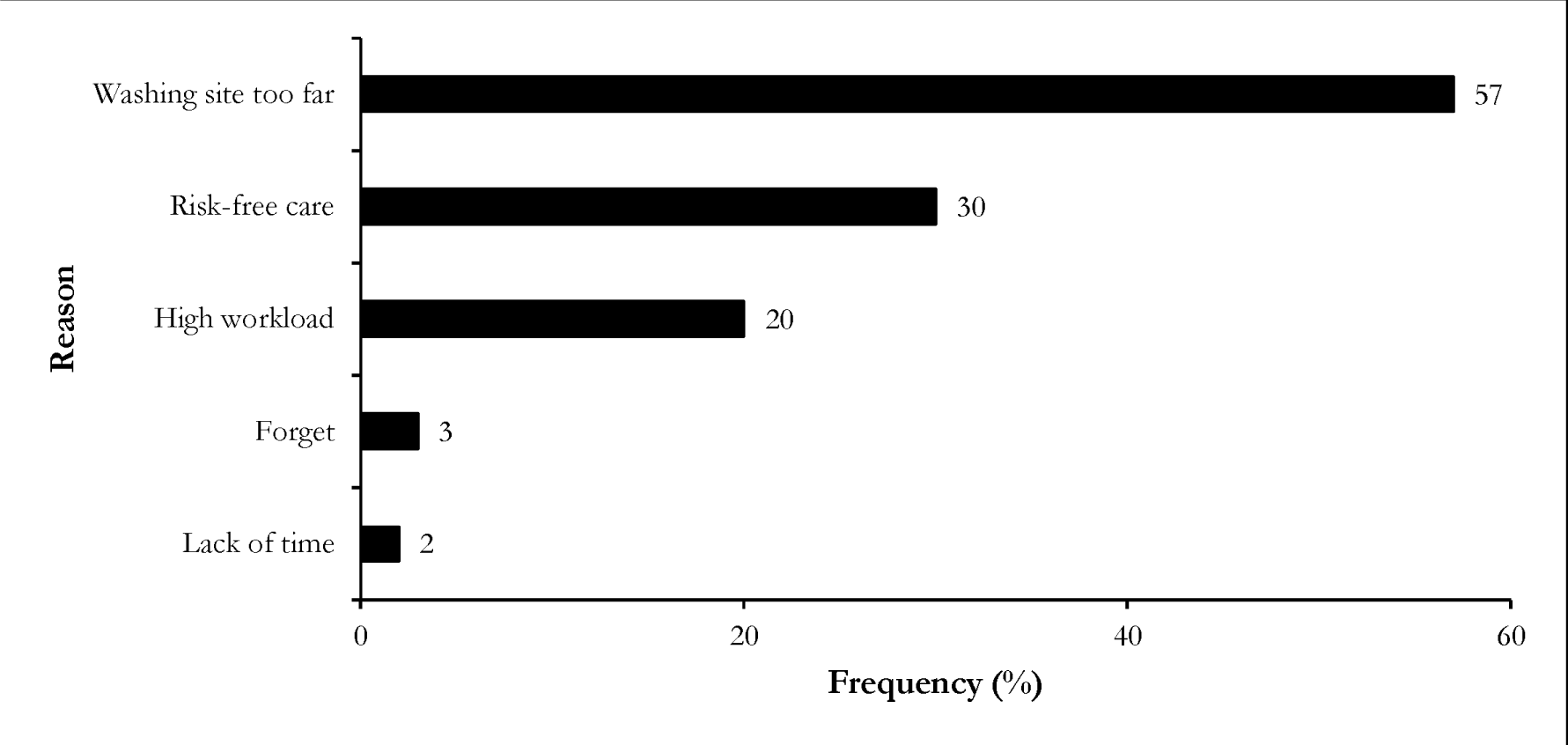
Constrains to systematic hand washing among healthcare workers (n=30)

Nearly one-third of HCWs reported (34%, n=14) having received training in ICP measures, with paramedics being the most represented (52%). In contrast, no medical students reported having received this training.

Most of HCWs reported having received hand hygiene training, with a particularly high proportion among medical students (80%). In addition, most of participants (80%) reported having received training two years or more ago (Table 2).

**Table 2.**
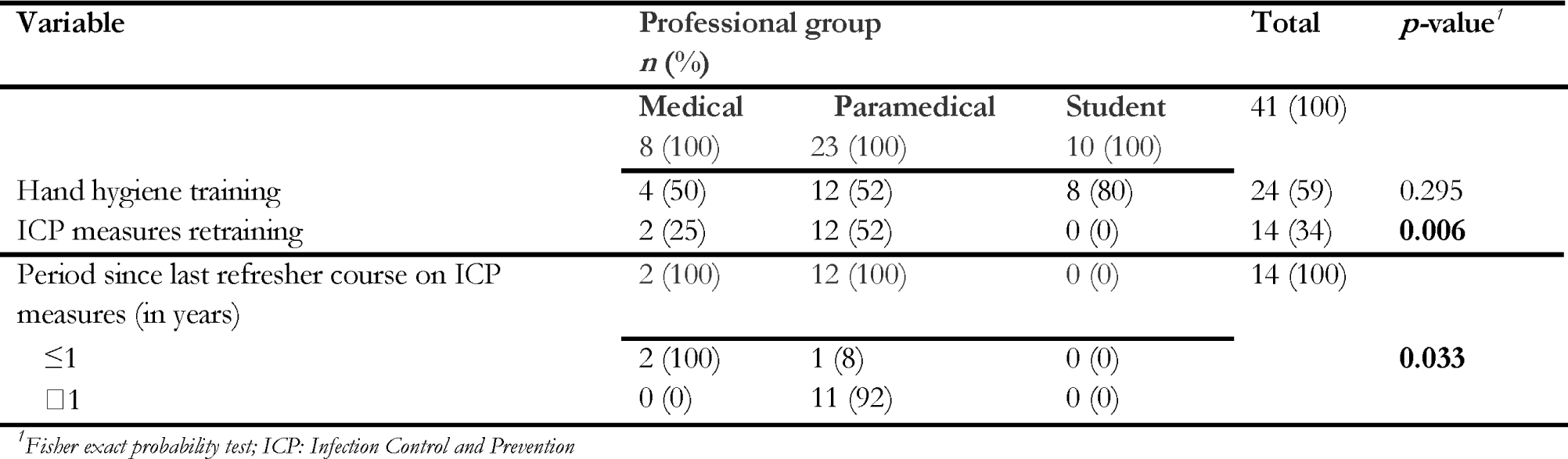
Reported follow-up of training sessions by professional group.

### Personal protective equipment

The most reported available hygiene devices included safety boxes (66%) and bleach for disinfection (78%). However, the equipment most frequently reported as unavailable was goggles/face shields (81%) and clean single-use towels (78%) (Figures 2 and 3).

**Fig. 2.**
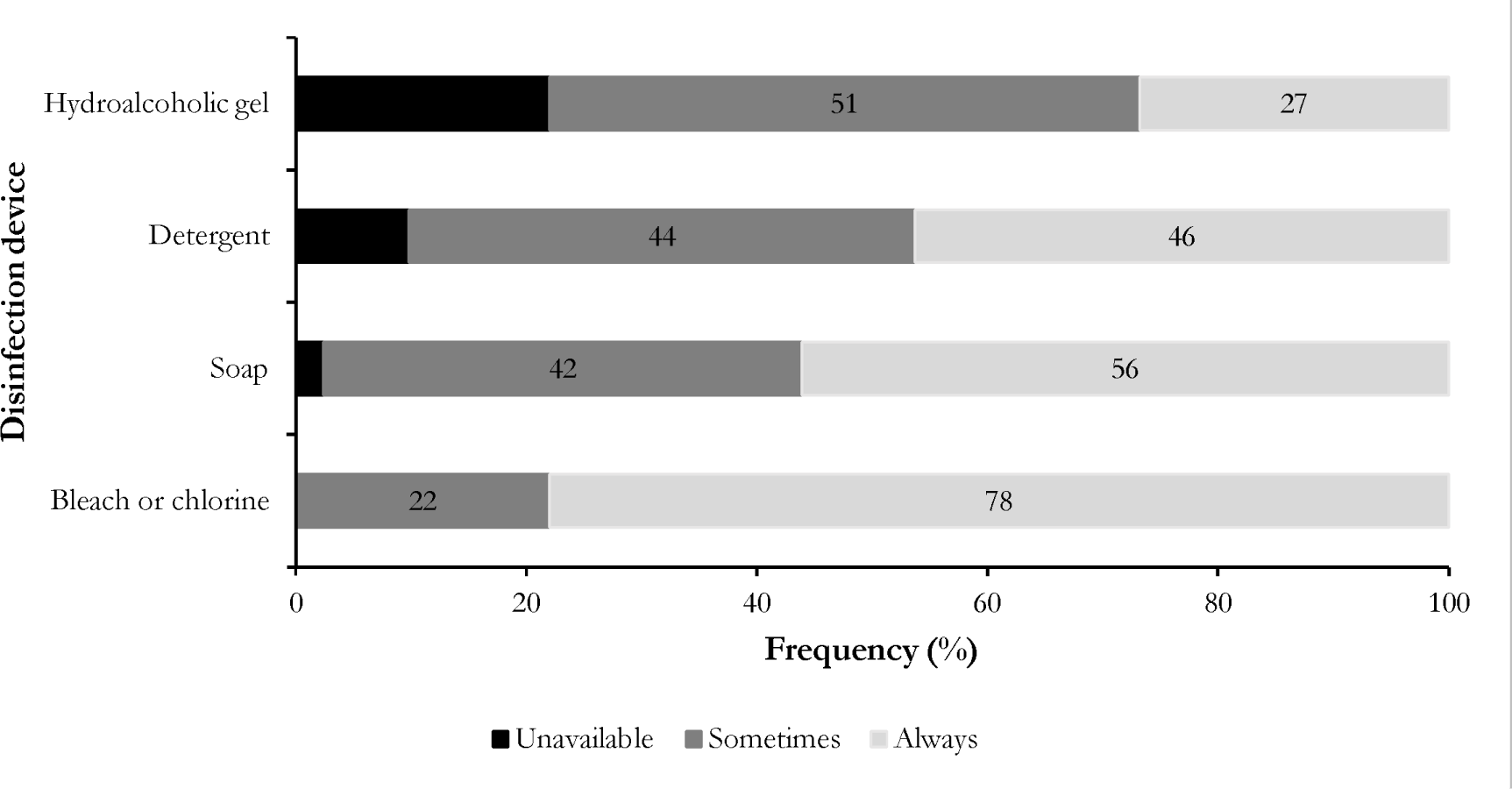
Perception of disinfectant availability among healthcare workers (n=41)

**Fig. 3.**
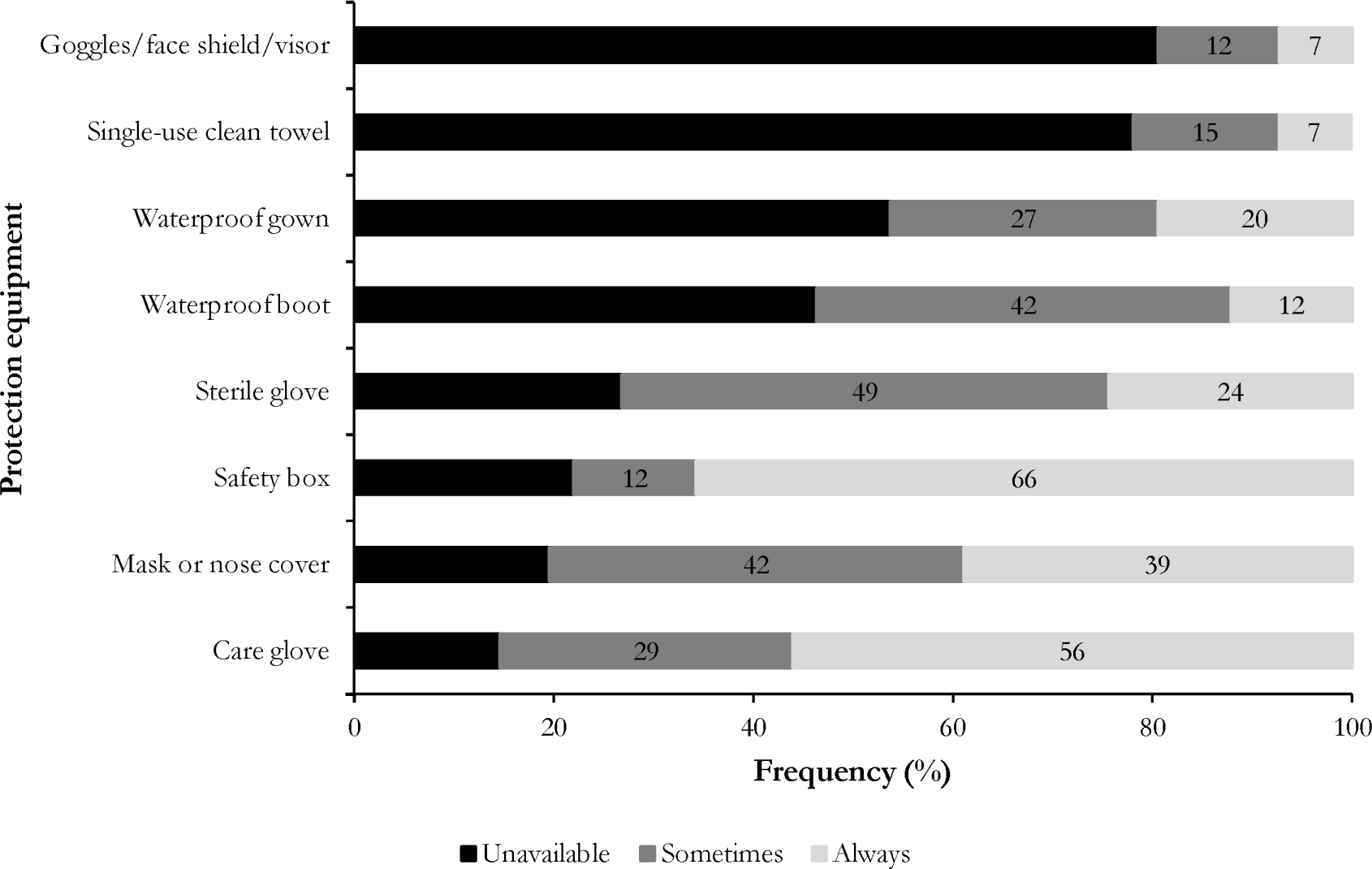
Perception of personal protective equipment availability among healthcare workers (n=41)

### Waste management and sterilization

Less than one-third of respondents (24%) reported the availability of posters on equipment sterilization protocols. More than half of HCWs (54%) reported having information on the correct preparation of a decontamination solution. There was no significant difference between different professional groups (Table 3).

**Table 3.** Perception by professional group of the availability of information brochures/poster on infection prevention and control.

| Variable | Professional group |  |  | Total | *p*-value<sup>†</sup> |
| --- | --- | --- | --- | --- | --- |
|  | *n* (%) |  |  |  |  |
|  | Medical | Paramedical | Student | 41 (100) |  |
|  | 8 (100) | 23 (100) | 10 (100) |  |  |
| Sterilization protocol displayed | 3 (38) | 4 (17) | 3 (30) | 10 (24) | 0,554 |
| Waste management guide available | 4 (50) | 14 (61) | 2 (20) | 20 (49) | 0,238 |
| Information on decontamination solution preparation available | 4 (50) | 14 (61) | 4 (40) | 22 (54) | 0,583 |

<sup>†</sup> Fisher exact probability test

### Framework for implementing infection control and prevention

The assessment of ICP measure implementation revealed that the maternity subunit was at an intermediate level, while the inpatient sub-unit was at a basic level. The overall score indicated that the service was at a basic level of ICP measure implementation. The highest scores were obtained in components related to workload (58.8%), guidelines (57.6%), and structural aspects of ICP (52.5%). In contrast, the lowest score (5%) was observed in the essential component evaluating multimodal strategies for ICP implementation (Table 4).

**Table 4.** Assessment of the implementation of the infection control and prevention framework.

| Essential Component | Sub-unit |  | Score |
| --- | --- | --- | --- |
|  | Maternity | Inpatient |  |
| Structural aspects of ICP | 95.0 | 10.0 | 52.5 |
| Guidelines for ICP | 44.4 | 70.8 | 57.6 |
| ICP education and training | 50.0 | 22.2 | 36.1 |
| Healthcare-associated infections surveillance | 41.0 | 10.3 | 25.6 |
| Multimodal strategies for implementing ICP interventions | 10.0 | 0.0 | 5.0 |
| Monitoring/auditing of serious injury prevention practices and feedback | 37.5 | 10.0 | 23.8 |
| Tasks, staffing and bed occupancy | 58.8 | 58.8 | 58.8 |
| Creation of environments and acquisition of care materials and equipment | 79.5 | 72.7 | 76.1 |
| **Total Score** | **416.3** | **254.9** | **335.6** |

ICP: Infection Control and Prevention

### Microbiological analysis of environmental surfaces and medical devices

Fifteen samples were collected from each sub-unit (inpatient and maternity). All 30 samples collected showed bacterial contamination. Fifty-five isolates were identified, most of which were Gram-positive (60%) and from the inpatient subunit and (62%). The tap was the most contaminated surface, with 14 identified isolates (Table 5).

**Table 5.** Identified isolates by Gram category and surfaces/medical devices.

| Characteristic | Gram Category<br>*n* (%) |  | Total | *p*-value<sup>†</sup> |
| --- | --- | --- | --- | --- |
|  | Negative | Positive | 55 (100) |  |
|  | 22 (40) | 33 (60) |  |  |
| **Sub-Unit** |  |  |  | 0.258 |
| Inpatient | 16 (47) | 18 (53) | 34 |  |
| Maternity | 6 (29) | 15 (71) | 21 |  |
| **Surface & Devices** |  |  |  | 0.723 |
| Trolley | 4 (40) | 6 (60) | 10 |  |
| Door handle | 3 (25) | 9 (75) | 12 |  |
| Drip stand | 4 (67) | 2 (33) | 6 |  |
| Tap | 6 (43) | 8 (57) | 14 |  |
| Delivery table | 2 (40) | 3 (60) | 5 |  |
| Nursing table | 3 (37) | 5 (63) | 8 |  |

<sup>†</sup>Fisher exact probability test

The most prevalent microbial strains in the department included Staphylococcus aureus (36%), other Staphylococcus spp. (24%), Proteus mirabilis (13%) and Klebsiella species (7%). Non-enterobacterial Gram-negative rods were the least represented (Figure 4).

**Fig. 4.**
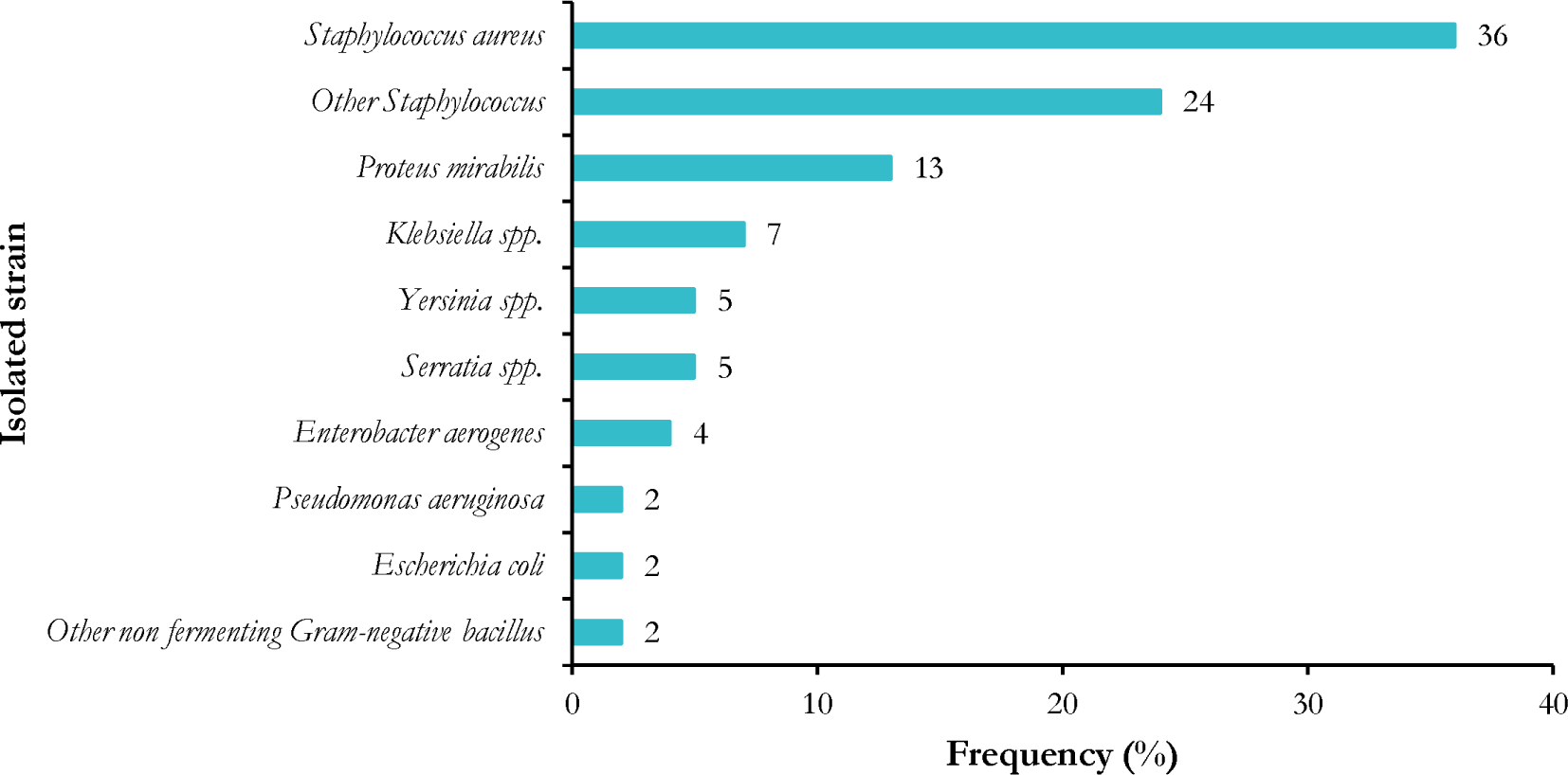
Profile of bacteria present on environmental surfaces and medical devices (n=55)

Most of microorganisms identified (70%) in the maternity ward belonged to the Staphylococcus spp., with S. aureus being the most represented species (42%). However, a nearly equivalent distribution was observed between Staphylococci (52%) and Enterobacteria (45%) (Table 6).

**Table 6.**
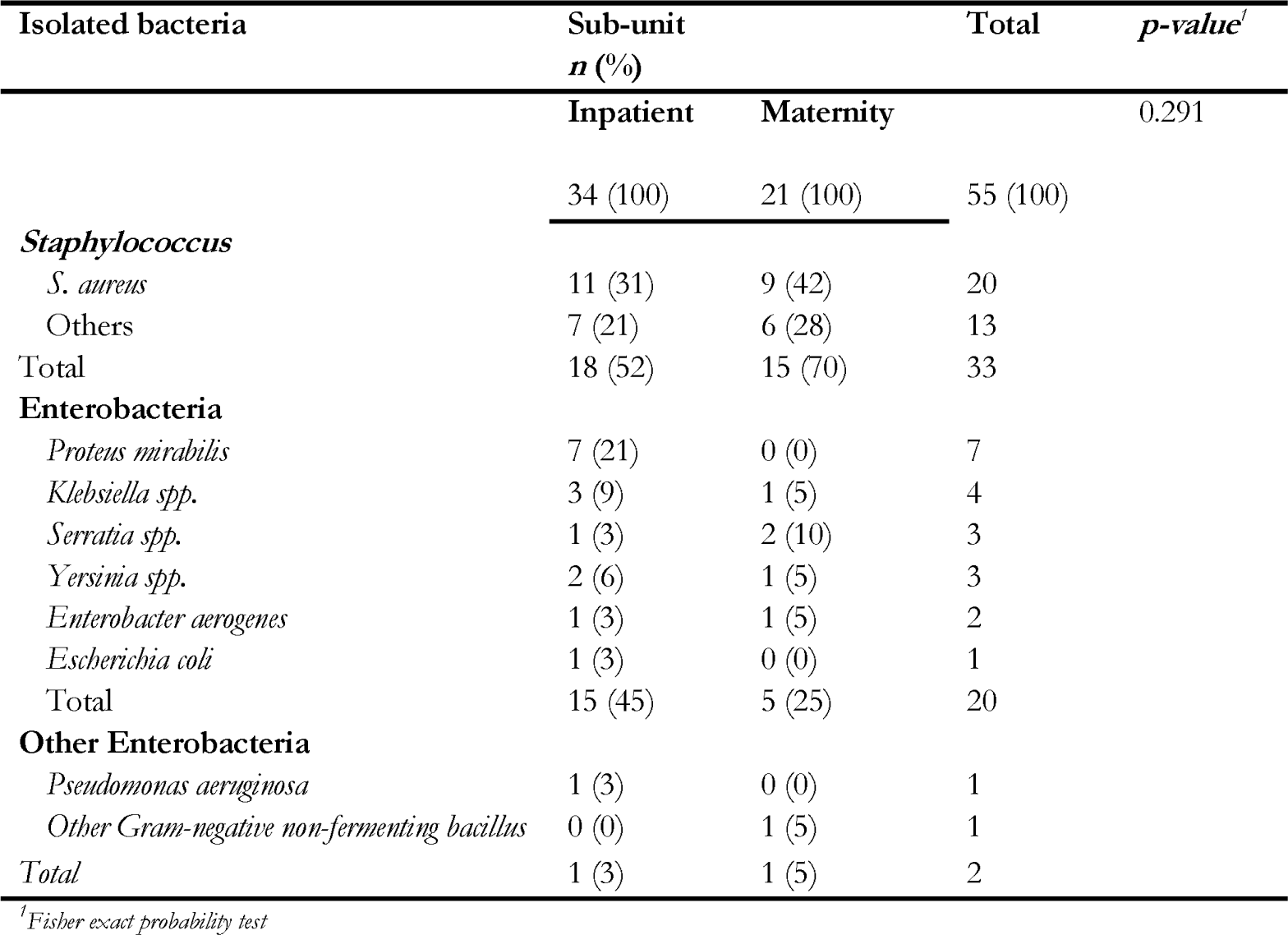
Bacterial strains isolated in the two sub-units.

Most microorganisms (74%) identified on door handles belonged to the Staphylococcus spp., with S. aureus being the most represented species (57%). In contrast, the trolley was primarily soiled by Enterobacteriaceae (66%), with the most isolated germs belonging to Proteus mirabilis (33%) and Klebsiella (33%) species (Table 6).

**Table 6.**
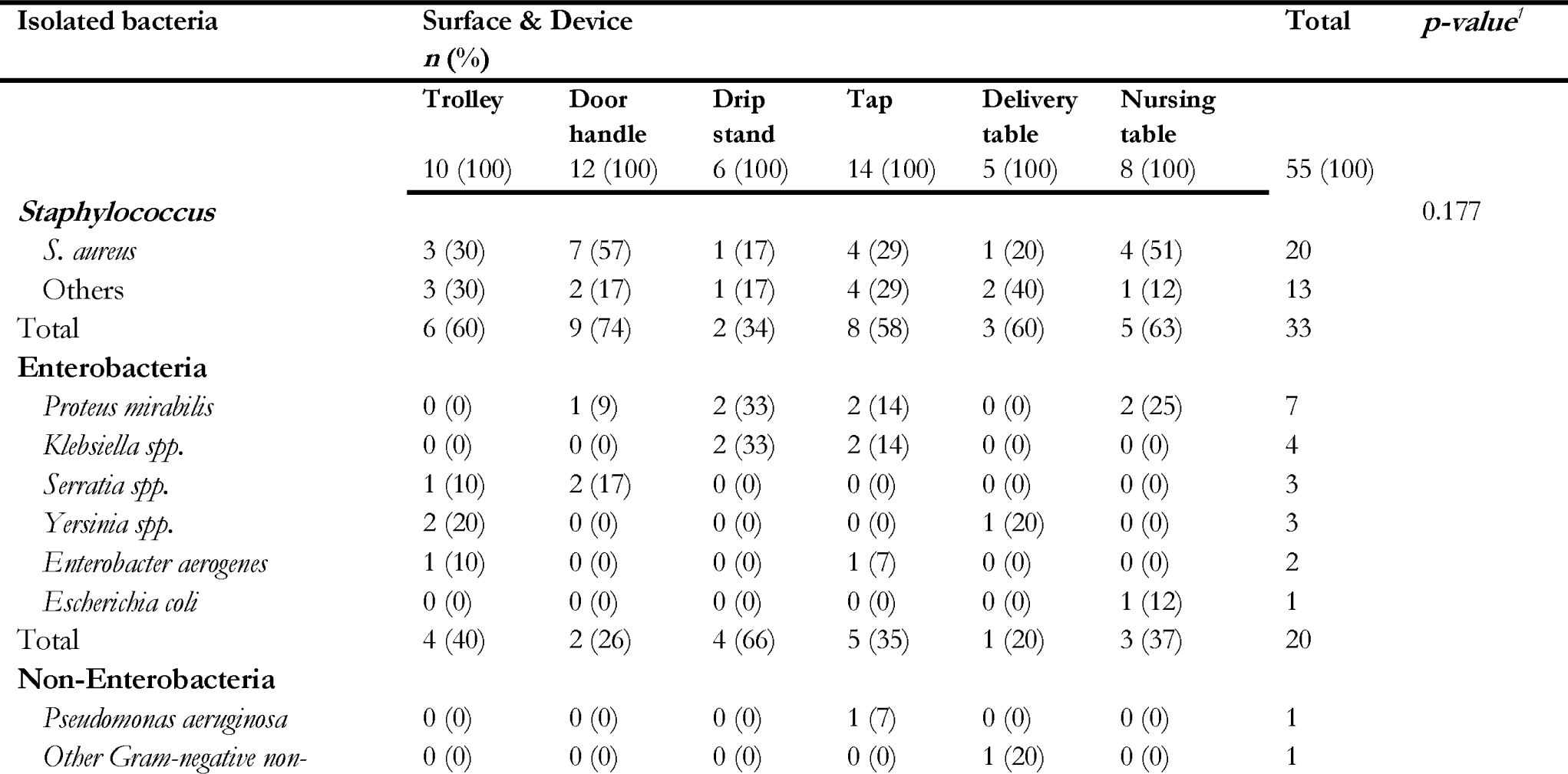
Classification of bacteria strains isolated from environmental surfaces and medical devices.

### Antibiotic susceptibility of isolated staphylococcal strains

Most staphylococci isolated were penicillinase producers (≥ 90%). Significant resistance was also observed for fosfomycin, rifampicin, fusidic acid and cotrimoxazole (≥ 70%). In contrast, the most effective antibiotics against these organisms were ciprofloxacin, erythromycin and vancomycin, which showed low or no resistance (Table 7).

**Table 7.** Antibiotic resistance profile of isolated staphylococci (n=33)

| Tested antibiotic | *Staphylococcus*<br>*n* (%) |  |
| --- | --- | --- |
|  | *S. aureus*<br>20 (100) | *Others*<br>13 (100) |
| **Penicillin** |  |  |
| Penicillin G | 20 (100) | 13 (100) |
| Ampicillin | 18 (90) | 12 (92) |
| Oxacillin | 18 (90) | 13 (100) |
| **Cephalosporin** |  |  |
| Cefoxitin | 6 (30) | 5 (38) |
| **Aminoglycoside** |  |  |
| Gentamicin | 3 (15) | 4 (31) |
| **Quinolone** |  |  |
| Ofloxacin | 3 (15) | 1 (8) |
| Ciprofloxacin | 1 (5) | 0 (0) |
| Norfloxacin | 1 (5) | 1 (8) |
| **Cycline** |  |  |
| Tetracycline | 8 (40) | 4 (31) |
| **Fusidanine** |  |  |
| Fusidic acid | 15 (75) | 9 (69) |
| **Glycopeptide** |  |  |
| Vancomycin | 1 (5) | 0 (0) |
| **Sulfonamide and Diamino-Pyrimidine** |  |  |
| Sulfamethoxazole + Trimethoprim | 15 (75) | 9 (69) |
| **Macrolide** |  |  |
| Erythromycin | 5 (25) | 0 (0) |
| **Streptogramin (synergistin)** |  |  |
| Pristinamycin | 4 (20) | 1 (8) |
| **Phosphonic acid** |  |  |
| Fosfomycin | 18 (90) | 13 (100) |
| **Rifamycin** |  |  |
| Rifampicin | 17 (85) | 13 (100) |

Methicillin resistance was observed in 70% of isolated Staphylococcus strains, with no significant difference between department sub-units (p=0.722). The primary contaminated medical devices were trolleys (100%), delivery tables (100%), and door handles (78%) (Table 8).

**Table 8.** Prevalence and distribution of methicillin-resistant staphylococci (n=33)

| Variable | Count ($n$) | Frequency (%) | Total | $p$-value<sup>†</sup> |
| --- | --- | --- | --- | --- |
| ***Staphylococcus*** |  |  |  | >0.999 |
| S. aureus | 14 | 70 | 20 |  |
| Others | 9 | 69 | 13 |  |
| **Sub-Unit** |  |  |  | 0.722 |
| Inpatient | 12 | 67 | 18 |  |
| Maternity | 11 | 73 | 15 |  |
| **Surface & Device** |  |  |  | 0.667 |
| Trolley | 3 | 50 | 6 |  |
| Door handle | 7 | 78 | 9 |  |
| Drip stand | 2 | 100 | 2 |  |
| Tap | 5 | 62 | 8 |  |
| Delivery table | 3 | 100 | 3 |  |
| Nursing table | 3 | 60 | 5 |  |

<sup>†</sup>Fisher exact probability test

### Antimicrobial susceptibility of gram-negative bacterial strains

Most identified Gram-negative bacteria produced penicillinases (≥70%) and cephalosporinases (≥80%), particularly against cefuroxime, cefepime, and cefoxitin. in contrast, cefotaxime and quinolone antibiotics (nalidixic acid, ciprofloxacin, and levofloxacin) were the most effective against these bacteria.

High resistance (near 100%) was observed among gram-negative bacteria for aztreonam, fosfomycin, and trimethoprim used alone. however, combination with sulfonamides (cotrimoxazole) overcame this resistance. The bacteria were highly susceptible to carbapenems (imipenem and meropenem) and vancomycin (Table 9)

**Table 9.** Resistance profile to standard antibiotics of Gram-negative bacilli circulating in the care unit (n=22)

| Tested antibiotic | *Enterobacteria*<br>n (%) |  |  |  |  |  | *Non-Enterobacteria*<br>n (%) |  |
| --- | --- | --- | --- | --- | --- | --- | --- | --- |
|  | *Proteus mirabilis* | *Klebsiella spp.* | *Yersinia spp.* | *Serratia spp.* | *Enterobacter aerogenes* | *Escherichia coli* | *Pseudomonas aeruginosa* | *Other Gram-negative non-fermenting bacillus* |
|  | 7 (100) | 4 (100) | 3 (100) | 3 (100) | 2 (100) | 1 (100) | 1 (100) | 1 (100) |
| **Penicillin** |  |  |  |  |  |  |  |  |
| Amoxicillin | 5 (71) | 3 (75) | 2 (67) | 3 (100) | 2 (100) | 0 (0) | 1 (100) | 1 (100) |
| Amoxicillin + Clavulanate | 5 (71) | 3 (75) | 2 (67) | 3 (100) | 2 (100) | 0 (0) | 1 (100) | 1 (100) |
| **Aminoglycoside** |  |  |  |  |  |  |  |  |
| Kanamycin | 3 (43) | 3 (75) | 3 (100) | 1 (33) | 0 (0) | 0 (0) | 1 (100) | 0 (0) |
| **Cephalosporin** |  |  |  |  |  |  |  |  |
| Ceftazidime | 4 (57) | 1 (25) | 2 (67) | 2 (67) | 1 (50) | 0 (0) | 1 (100) | 0 (0) |
| Cefuroxime | 7 (100) | 4 (100) | 3 (100) | 3 (100) | 2 (100) | 1 (100) | 1 (100) | 1 (100) |
| Cefepime | 6 (86) | 4 (100) | 3 (100) | 3 (100) | 2 (100) | 0 (0) | 1 (100) | 1 (100) |
| Cefotaxime | 0 (0) | 0 (0) | 0 (0) | 0 (0) | 0 (0) | 0 (0) | 0 (0) | 0 (0) |
| Ceftriaxone | 2 (29) | 3 (75) | 2 (67) | 3 (100) | 2 (100) | 1 (100) | 0 (0) | 0 (0) |
| Cefoxitin | 6 (86) | 4 (100) | 3 (100) | 3 (100) | 1 (50) | 1 (100) | 1 (100) | 1 (100) |
| **Quinolone** |  |  |  |  |  |  |  |  |
| Nalidixic acid | 0 (0) | 0 (0) | 0 (0) | 0 (0) | 1 (50) | 0 (0) | 0 (0) | 0 (0) |
| Ciprofloxacin | 0 (0) | 0 (0) | 0 (0) | 0 (0) | 0 (0) | 0 (0) | 0 (0) | 0 (0) |
| Levofloxacin | 0 (0) | 0 (0) | 0 (0) | 0 (0) | 0 (0) | 0 (0) | 0 (0) | 0 (0) |
| **Carbapenem** |  |  |  |  |  |  |  |  |
| Meropenem | 0 (0) | 0 (0) | 0 (0) | 1 (33) | 0 (0) | 0 (0) | 0 (0) | 1 (100) |
| Imipenem | 0 (0) | 0 (0) | 0 (0) | 0 (0) | 0 (0) | 0 (0) | 0 (0) | 0 (0) |
| **Glycopeptide** |  |  |  |  |  |  |  |  |
| Vancomycin | 0 (0) | 0 (0) | 0 (0) | 0 (0) | 0 (0) | 0 (0) | 0 (0) | 0 (0) |
| **Phosphonic acid** |  |  |  |  |  |  |  |  |
| Fosfomycin | 6 (86) | 3 (75) | 3 (100) | 2 (67) | 2 (100) | 1 (100) | 1 (100) | 1 (100) |
| **Diamino-Pyrimidine** |  |  |  |  |  |  |  |  |
| Trimethoprim | 7 (100) | 3 (75) | 2 (67) | 2 (67) | 2 (100) | 1 (100) | 1 (100) | 1 (100) |
| **Sulfonamide and Diamino-Pyrimidine** |  |  |  |  |  |  |  |  |
| Sulfamethoxazole + Trimethoprim | 1 (14) | 0 (0) | 1 (33) | 0 (0) | 0 (0) | 0 (0) | 1 (100) | 0 (0) |
| **Monobactam** |  |  |  |  |  |  |  |  |
| Aztreonam | 4 (57) | 3 (75) | 2 (67) | 3 (100) | 1 (50) | 0 (0) | 1 (100) | 1 (100) |

### Multidrug resistance assessment

Multidrug resistance was observed in 100% of the bacteria isolated from the inpatient sub-unit.

Nearly all environmental surfaces and medical devices harbored multidrug-resistant bacteria (Table 11).

**Table 11.** Phenotype and location of multidrug-resistant bacteria (n=55)

| Variable | Characteristic | Count ($n$) | Frequency (%) | Total | $p$-value<sup>†</sup> |
| --- | --- | --- | --- | --- | --- |
| Location | **Sub-Unit** |  |  |  | 0.051 |
|  | Inpatient | 34 | 100 | 34 |  |
|  | Maternity | 18 | 86 | 21 |  |
|  | **Surface & Device** |  |  |  | 0.725 |
|  | Tap | 13 | 93 | 14 |  |
|  | Trolley | 10 | 100 | 10 |  |
|  | Door handle | 10 | 83 | 12 |  |
|  | Nursing table | 8 | 100 | 8 |  |
|  | Drip stand | 6 | 100 | 6 |  |
| Phenotype | **Staphylococcus** |  |  |  | 0.726 |
|  | *S. aureus* | 17 | 85 | 20 |  |
|  | Others | 13 | 100 | 13 |  |
|  | **Enterobacteria** |  |  |  |  |
|  | *Proteus mirabilis* | 7 | 100 | 7 |  |
|  | *Klebsiella* spp. | 4 | 100 | 4 |  |
|  | *Yersinia* spp. | 3 | 100 | 3 |  |
|  | *Serratia* spp. | 3 | 100 | 3 |  |
|  | *Enterobacter aerogenes* | 2 | 100 | 2 |  |
|  | *Escherichia coli* | 1 | 100 | 1 |  |
|  | **Non-Enterobacteria** |  |  |  |  |
|  | *Pseudomonas aeruginosa* | 1 | 100 | 1 |  |
|  | Other non-fermenting | 1 | 100 | 1 |  |
|  | Gram-negative bacillus |  |  |  |  |

<sup>†</sup>Fisher exact probability test

## Discussion

Healthcare-associated infections (HAIs) represent a significant threat to patient safety, resulting in serious complications, additional costs, and increased mortality [5]. These infections affect a substantial number of admitted patients, underscoring the need for effective prevention and control measures [3]. The One Health approach emphasizes the role of the environment in the development and transmission of infectious agents, including emerging and re-emerging pathogens. In Cameroon, maternal and neonatal mortality rates remain alarmingly high [8,9]. To address this, the country has committed to implementing universal health coverage [30–32].

### Knowledge of hand hygiene

Our study revealed alarmingly poor knowledge among midwives (29%) and younger participants, especially medical students (30%). Notably, those who reported receiving specific hand hygiene training did not demonstrate satisfactory knowledge of this topic. Given that most of those trained were medical students (80%), this raises concerns about their willingness to retain information about hygiene practices to ensure safe care. This highlights the need for refresher training on ICP, particularly hand hygiene, before clinical internships begin. In fact, no student reported recent refresher training on this topic. In addition, 80% of respondents reported that their last hand hygiene training was two or more years ago. These knowledge gaps contribute to poor practice, putting both patients and HCWs at risk of acquiring multidrug-resistant hospital-acquired pathogens. Therefore, implementing ongoing education and training programs for HCWs, especially medical students, is critical to improving their hand hygiene knowledge and skills and ultimately reducing the risk of HAIs.

### Personal protective equipment

The most commonly reported unavailable items were goggles/face shields and clean disposable towels. Studies conducted in Buea, Bertoua, and Yaoundé reported similar proportions in both primary and secondary referral hospitals [33–35]. These shortages are concerning, given that HCWs in maternity wards are regularly exposed to splashes on the mucous membranes of the face during childbirth [36–38]. These findings suggest that health facilities need to intensify their efforts to ensure the availability and accessibility of hygiene and personal protection equipment. This may include the procurement and regular maintenance of equipment, as well as comprehensive staff training on the importance of hygiene and personal protection. Finally, these findings highlight the need for regular monitoring and evaluation of hygiene and personal protection practices in health facilities to identify gaps and areas for improvement [36,39].

### Infection prevention and control assessment framework

The overall score indicated that the ward was at a basic level of implementation of ICP measures. This suggests that while some key elements of infection control are in place, their implementation remains inadequate. Additional efforts are necessary to strengthen these components and ensure effective infection control. This score was similar to that of the third reference hospital in Bangladesh (355.0 [Q1-Q3: 252.5 - 397.5] out of 800) and was consistent with the results of assessments reported in global analyses, which found the lowest scores (100 - 450 out of 800) in low- and middle-income countries [24,40]. The lowest scores were recorded in the core components assessing ICP education and training (36.1%), surveillance of HAIs (25.6%), multimodal strategies for implementing ICP interventions (5%), and monitoring/auditing of serious injury prevention practices and feedback (23.8%). These results corroborated observations made in 68 referral hospitals in Turkey and were lower than those obtained in 1442 referral hospitals in the Republic of Korea, likely because previous legislative and multimodal measures had been implemented in most of the hospitals surveyed [41,42]. This finding highlights a significant gap in the education and training of HCWs in ICP. A lack of knowledge can lead to poor practice and medical errors that are detrimental to both patient health and HCW safety [43–45].

### Microbial flora profile of surfaces and equipment

Most of the isolates identified originated from the inpatient unit, which can be attributed to the greater interaction among HCWs, patients, their families, and the healthcare environment compared to the maternity unit, where access is restricted to HCWs and patients. Furthermore, the implementation of ICP measures such as hygiene sessions and regular cleaning of medical equipment in the maternity unit could also contribute to these results.

The tap was found to be the most contaminated surface, while the trolley exhibited the highest contamination rate relative to the number of samples analyzed. Hand-washing areas, frequently used by HCWs for hand and equipment hygiene, are prime sites for bacterial colonization, particularly in the absence of regular and appropriate cleaning by cleaning staff. Bacterial contamination of these areas can seriously compromise hand hygiene and facilitate the transmission of pathogens to patients. Similarly, trolleys used to transport drugs and nursing equipment can act as vehicles for the transmission of germs from one patient to another if hand hygiene precautions such as hand washing, wearing gloves, and the use of hydro-alcoholic gel are not systematically followed. Our findings corroborate observations made at Treichville University Hospital in Côte d’Ivoire and the Centre Médical le Jourdain in Cameroon, where wash stations and trolleys were also the most contaminated sites [16,26].

The most common microbial strains in the department belonged to the species Staphylococcus aureus. These bacteria have developed a remarkable ability to adapt to the healthcare environment and their human host. S. aureus is part of the commensal microbiota of the nasal mucosa in approximately 20 to 40% of the general population but is also responsible for the transient colonization of the nasal mucosa in at least 60% of the remaining population. This may explain its predominance in this study [46,47]. It is the most clinically relevant staphylococcal species due to its involvement in HAIs and is the second most common pathogen responsible for this type of infection after E. coli [47]. S. aureus infects humans when skin and mucosal barriers are disrupted, for example, as a result of chronic skin conditions, wounds, or surgical procedures, allowing access to underlying tissues or the bloodstream and causing infection. Individuals with invasive medical devices or weakened immune systems, including pregnant women and premature newborns, are particularly susceptible to S. aureus infection [46–48]. Study reports have identified this bacterium as responsible for HAIs of the skin and soft tissues at Yaoundé University Teaching Hospital [19]. Our results were similar to those obtained at the Douala General Hospital, where S. aureus was the most frequently isolated bacterium [18].

Proteus mirabilis and Klebsiella spp. were the main pathogenic enterobacteria isolated in the department. These infectious agents are responsible for HAIs, especially in obstetrics-gynecology wards [3,19,49]. Proteus is found in abundance in soil and water and, although part of the normal human intestinal flora [49–51]. It is frequently responsible for infections of the human urinary tract, where it causes urinary tract infections and catheter-associated infections [52]. This is of concern given that the obstetrics-gynecology department regularly performs cesarean sections requiring the use of urinary catheters in parturient women. The circulation of this pathogen in the department increases the risk of women developing iatrogenic urinary tract infections after delivery [51,53].

Bacteria from Klebsiella spp. are responsible for infections characterized by high morbidity and mortality and by their ability to metastasize within the infected organism to cause sepsis. Risk factors for this infection include local healthcare practices, antibiotic misuse, and compliance with infection control procedures [54,55]. Poor compliance with standard precautions in the obstetrics-gynecology department could lead to contamination, as this bacterium has also been implicated in severe neonatal sepsis [56]. In addition, K. pneumoniae is one of the most commonly isolated pathogens in healthcare settings and is responsible for HAIs in Cameroon, Côte d’Ivoire, and Mali [3,16–19].

### Susceptibility profile of isolated staphylococcal strains

Antimicrobial resistance analysis revealed that most staphylococci isolates were penicillinase producers (≥ 90%). In addition, significant resistance rates were observed for fosfomycin, rifampicin, fusidic acid, and cotrimoxazole (≥ 70%).

This proportion of penicillin-resistant strains was similar to that observed in studies conducted in Cameroon and higher than that found at the University Hospital of Treichville, Côte d’Ivoire (28%) [16,17,39]. This difference could indicate a less rational use of antibiotics in Cameroon, which explains the resistance to common antibiotics, thereby limiting their effectiveness in the treatment of staphylococcal infections. Penicillin resistance is a major public health problem because it complicates the treatment of infections and increases the risk of complications and mortality [57].

Most Staphylococcus strains isolated (70%) were resistant to methicillin. Methicillin-resistant Staphylococcus aureus (MRSA) is a major cause of morbidity and mortality in neonates admitted to neonatal intensive care units. Neonatal MRSA colonization is attributed to various sources, including mothers, HCWs, and environmental surfaces. It can lead to life-threatening infections, prolonged hospital stays, and significant economic costs [48,58].

A meta-analysis of the worldwide distribution of MRSA revealed varying prevalence rates depending on the populations studied, ranging from 0.3 to 5.1% for mothers of newborns, from 3.1 to 18.4% for HCWs, with the highest rates (3.5 to 36.0%) observed in environmental samples, confirming the results of our study [58]. In addition, MRSA is a common cause of HAIs in many countries, including Cameroon and Côte d’Ivoire, where methicillin-resistant strains have been identified in 71-81% of staphylococci isolated from hospitals [16,17,59]. It is therefore essential to monitor antibiotic resistance and develop new strategies to combat penicillin-resistant staphylococcal infections.

### Susceptibility profile of Gram-negative bacterial strains

Most of the Gram-negative bacteria identified were penicillinase and cephalosporinase producers. A similar bacterial resistance profile was observed at the Centre Médical le Jourdain in Yaoundé [16].

The majority of the bacteria identified (85-100%) exhibited multidrug resistance. This remains a significant concern for the vulnerable patients in obstetrics-gynecology departments, particularly pregnant women, women in the immediate postpartum period, and their newborns. A meta-analysis conducted in Cameroon on human and environmental samples showed lower rates of multidrug resistance compared to those observed in our study [60,61]. The specific characteristics of the samples analyzed in our study could potentially explain these differences.

### Focus on multidrug resistance to antibiotics

Resistance to the most commonly prescribed antimicrobials leads to a significant increase in mortality, length of hospital stays, and healthcare costs. Faced with these multidrug-resistant infections and delays in obtaining microbiological results, broad-spectrum antibiotics are often used as empirical therapy. This practice can lead to the overuse and misuse of antibiotics, contributing to the development of resistance [62].

The high prevalence of multidrug-resistant bacterial strains underscores the existence of facilitating factors. These factors include poor compliance with regulations and a lack of control over the sale and use of antibiotics by HCWs and the general public [63,64]. In addition, the scarcity of accurate epidemiological data on antibiotic use and bacterial resistance in Cameroon complicates the development of effective strategies [15,65].

## Limitations

It is important to recognize that the evaluation model used was tailored to a specific health setting. Consequently, the results obtained may not accurately reflect the situation in the whole health facility. In addition, the sample size did not provide sufficient power to detect statistical significance for certain variables that were not considered significant. These limitations highlight the need for a more comprehensive study with a larger sample size.

## Conclusions

This study highlights significant risks that increase the likelihood of HAIs in neonates and mothers on the unit. Such deficiencies included inadequate staff knowledge of hand hygiene, unsystematic compliance with proper hand hygiene, inadequate availability of certain essential PPE and limited implementation of ICP measures by the unit, which was rated at a basic level of implementation. The presence of pathogenic and multi-resistant bacteria in the healthcare environment underscores the need to not only improve compliance with ICP measures but also develop a national policy to regulate the use of antibiotics, promote their rational use, build the capacity of HCWs to diagnose and treat infections appropriately, and finally to establish programs to monitor bacterial resistance and antibiotic use to track changes in the situation.

## Abbreviations

HAI: Healthcare Associated Infection HCW: Healthcare Worker
ICP: Infection Control and Prevention

## Declaration

Authors’ Contribution: Drafting of the study protocol, data collection, analysis and interpretation: F.Z.L.C.; Drafting and editing of manuscript: F.Z.L.C.; Critical revision of the manuscript: E.E.L.; Conception, design and supervision of research protocol and implementation, data analysis plan, revision, editing and final validation of the manuscript: I.T.

## Ethical Approval Statement

The protocol was approved by Institutional Review Board (IRB) of the Faculty of Medicine and Biomedical Sciences of Yaoundé 1 and the ethical clearance: N°1017/UYI/FMSB/VDRC/DAASR/CSD issued. Informed consent was obtained from participants prior to inclusion in the study. All methods were performed according to relevant guidelines and regulations.

## Consent for publication

Not applicable.

## Availability of data and materials

All data generated or analyzed during this study are included in this published article.

## Data Availability

All data produced in the present work are contained in the manuscript

## Acknowledgements

Our gratitude goes to the health staff who agreed to participate in this study and to the manager of the referral hospital general manager who gave an authorization to conduct the study.

